# Mathematical modelling projections versus the actual course of the COVID-19 epidemic following the nationwide lockdown in Kyrgyzstan

**DOI:** 10.1101/2020.12.10.20247247

**Authors:** Ainura Moldokmatova, Aida Estebesova, Aizhan Dooronbekova, Chynar Zhumalieva, Aibek Mukambetov, Talant Abdyldaev, Aisuluu Kubatova, Shamil Ibragimov, Nurbolot Usenbaev, Ainura Kutmanova, Lisa J White

## Abstract

Kyrgyzstan was placed under a two-month, nationwide lockdown due to the COVID-19 epidemic, starting on March 25, 2020. Given the highly disruptive effects of the lockdown on the national economy and people’s lives, the government decided not to extend lockdown beyond the initially planned date of May 10, 2020. The strategy chosen by the government was close to the input parameters of our model’s baseline scenario, ‘full lockdown release’, which we presented to policymakers in April 2020, along with various other hypothetical scenarios with managed lockdown release options. To explore whether our model could accurately predict the actual course of the epidemic following the release of lockdown, we compared the outputs of the baseline scenario, such as new cases, deaths, and demand for and occupancy of hospital beds, with actual official reports. Our analysis revealed that the model could accurately predict the timing of the epidemic peak, with a difference of just two weeks, although the magnitude of the peak was overestimated compared with the official statistics. However, it is important to note that the accuracy of the official reports remains debatable, so outputs relating to the size of the epidemic and related pressures on the health system will need to be updated if new evidence becomes available.

## Introduction

The first imported cases of COVID-19 in Kyrgyzstan were reported on March 16, 2020, followed by the declaration of a state of emergency and a nationwide, two-month, full ‘lockdown’, beginning on March 25, 2020. As part of the lockdown, the public health response in the country was focussed on non-pharmaceutical interventions, which included contact tracing, isolation of infected people and quarantining those who were exposed to infection, hand hygiene, physical distancing, a travel ban, and the closure of schools, offices, markets and other public spaces.

The lockdown helped Kyrgyzstan to effectively control the epidemic, during which the cumulative number of confirmed cases reached 1038, with 13 reported deaths [1]. However, the lockdown was associated with substantial social and economic disruption and led to public criticism of the government for taking such strict measures for such a long period of time. Under increasing public pressure, the government made the decision not to prolong the lockdown after the initially planned two-month period and considered options for other measures, balancing their effectiveness at reducing the transmission of COVID-19 with their impact on the societal and economic aspects of people’s lives.

Owing to the lack of knowledge and evidence around effective ways to prevent and treat COVID-19 in the local context and a rapidly developing pandemic globally, the examination of ‘what if’ scenarios through mathematical modelling became useful for providing important insights for public health decision-makers. To assist with this process, our team, an independent Kyrgyz modelling group, in collaboration with and receiving technical support from the international COVID-19 Modelling (CoMo) Consortium [2], reviewed several hypothetical lockdown-release scenarios. In April, 2020, we presented our findings to key decision-makers in Kyrgyzstan, including the Ministry of Health (MoH) and the National COVID-19 Response Unit (NCRU). We modelled the so-called baseline scenario, with full lockdown release, which was then compared with other hypothetical scenarios of managed lockdown release of various durations and intensities of post-lockdown measures. The details can be found in the Policy Notes, which we shared with decision-makers at the end of April 2020 (S1 Appendix).

On May 10, 2020, the Kyrgyzstan government made the decision to release the lockdown but retain a partial travel ban, case tracing, and continued school closures until the end of May. A few weeks later, the number of symptomatic COVID-19 cases increased tremendously, causing a significant burden on the health system. As the epidemic developed, hospitals began to experience a scarcity of hospital surge and intensive care unit (ICU) beds, oxygen ventilators, and human and other resources. As a result, many patients with severe COVID-19 symptoms were unable to access adequate hospital and ICU/oxygen treatment, and this contributed to the increasing number of deaths during the peak of the epidemic in July 2020.

In this paper, we analyse what our model was able to predict of the actual course of the epidemic and how close this prediction was to reality. In particular, our interest was focussed on the following two questions:

1. How accurately did the model predict the actual course of the epidemic?
2. How accurately did the model predict the actual hospital demand and occupancy and their effect on mortality?

## Methods

We applied the web-based interface of a dynamic SEIRS (susceptible–exposed–infected– recovered–susceptible) age-structured model for the COVID-19 pandemic, developed by the CoMo Consortium in collaboration with the Oxford Modelling for Global Health (OMGH) Group, for examining the effect of various intervention packages on the epidemic curve in each of more than 150 countries [3].

At the request of the Kyrgyzstan MoH, we sought effective and feasible post-lockdown intervention strategies that would help the country to control the epidemic, keep the number of severe cases at a reasonable level and prevent the health system from becoming overwhelmed during the epidemic peak. We reviewed five hypothetical scenarios for lockdown release: 1) baseline, full release; 2) managed lower intensity release; 3) managed higher intensity release; 4) prolonged lockdown with full release; and 5) prolonged lockdown with managed release.

As shown in Table 1, the ‘full lockdown release’ scenario implied there were no interventions other than hand hygiene and mask wearing once the 2-month lockdown was lifted. It should be noted that hand hygiene and mask wearing were included in this scenario with a comparatively low coverage, assuming that some of the population would continue following these two measures. In addition, a standard school closure period for summer holidays, from June to August, was taken into consideration, as in the other hypothetical scenarios. All other scenarios included either managed 2-month lockdown release options or extended lockdowns for an additional one or two months with full or managed release.

**Table 1.**
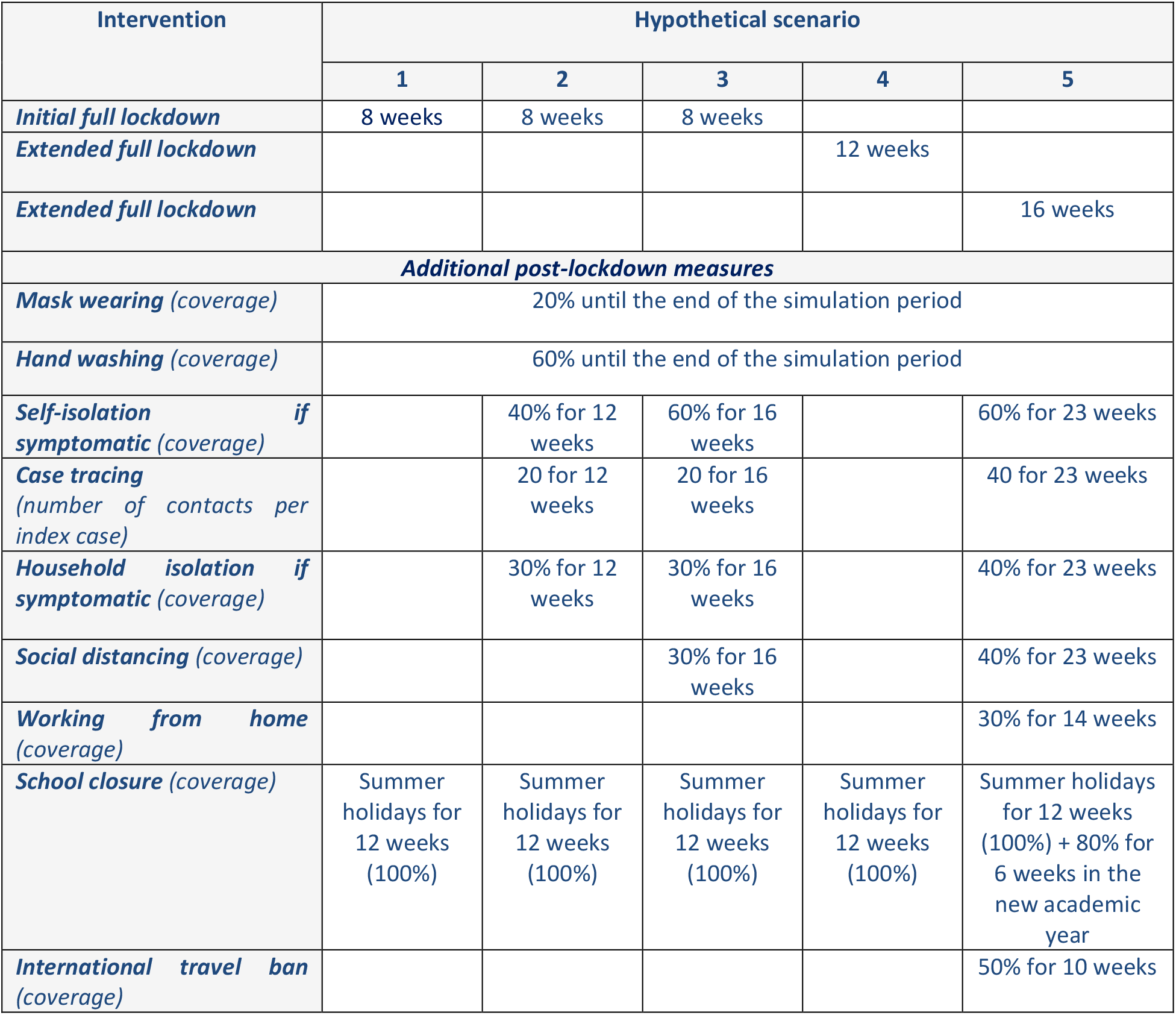
Intervention parameters for the hypothetical scenarios modelled.

Of the five options modelled, the strategy chosen by the government most closely resembled the input data of the full lockdown release scenario. As shown in Figure 1, the government made the decision to resume the normal mode of economic and social life of the country after May 10, 2020, continuing the school closure until the end of the academic year (May 30) and keeping a partial travel ban for an additional few weeks. In addition, the government continued case tracing; however, it was not feasible to adequately implement this strategy due to the increase in new cases with undefined contacts during the second part of June on the one hand and the shortage of human and other resources on the other hand. Accordingly, from July 3, 2020, the MoH stopped reporting the daily statistics of defined index contacts [4].

**Figure 1.**
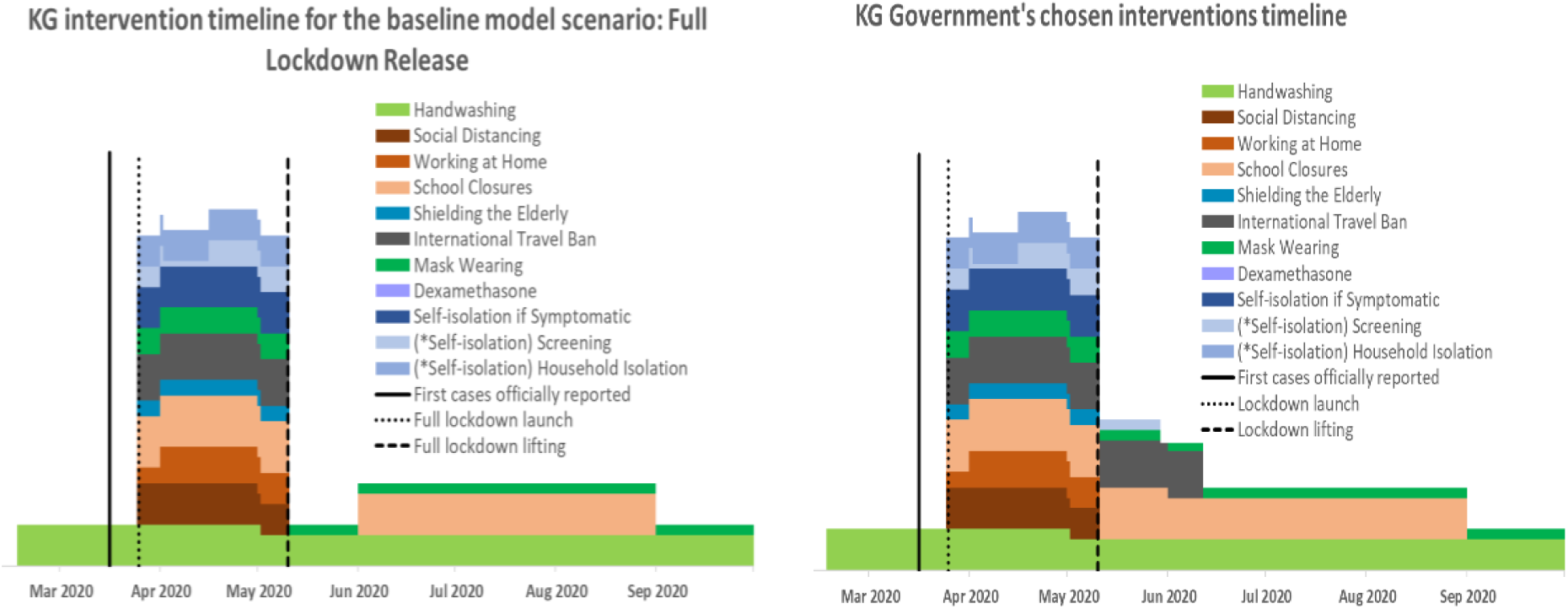
Timelines of interventions for the hypothetical ‘full lockdown release’ scenario and the government’s chosen strategy to release the lockdown on May 10, 2020. **Tool resource**: CoMo Consortium, 2020.

Evidence suggests that screening without isolation of positive cases and their contacts is less effective at controlling the epidemic than when such isolation is achieved [5,6]. However, in the Kyrgyz context it was not possible to take this measure due the local social and cultural norms, such as the extended intergenerational household structure of the majority of the population, as well as strong family and tribal networks and related large social gatherings.

Based on the above, we compared the model output data for the full lockdown release scenario with the actual course of the epidemic in Kyrgyzstan, following the release of lockdown on May 10, 2020. It is important to note that the model was visually fitted against actual new cases and deaths up to April 24, 2020, as part of the simulation procedure in the web-based interface. The charts of the visual calibration outputs with key model parameters can be found in S2 Appendix, Parts I and II.

## Results

### How accurately did the model predict the epidemic for the full lockdown release scenario?

The majority of COVID-19 cases are asymptomatic or have mild symptoms, particularly among younger people [7–9]. Considering the population structure and limited testing capacity in Kyrgyzstan, our model predicted a significantly higher number of unreported asymptomatic cases or cases with mild symptoms compared with the officially reported number of cases. According to the model, a full lockdown release would be followed by an intense increase in new cases within the next few weeks. The peak was predicted to occur somewhere between the end of June and the first half of July, with approximately 14,000 reported cases and 180,000 unreported cases per day expected to be observed during the peak of the epidemic, if the lockdown was lifted as planned on May 10 (Figure 2A).

**Figure 2.**
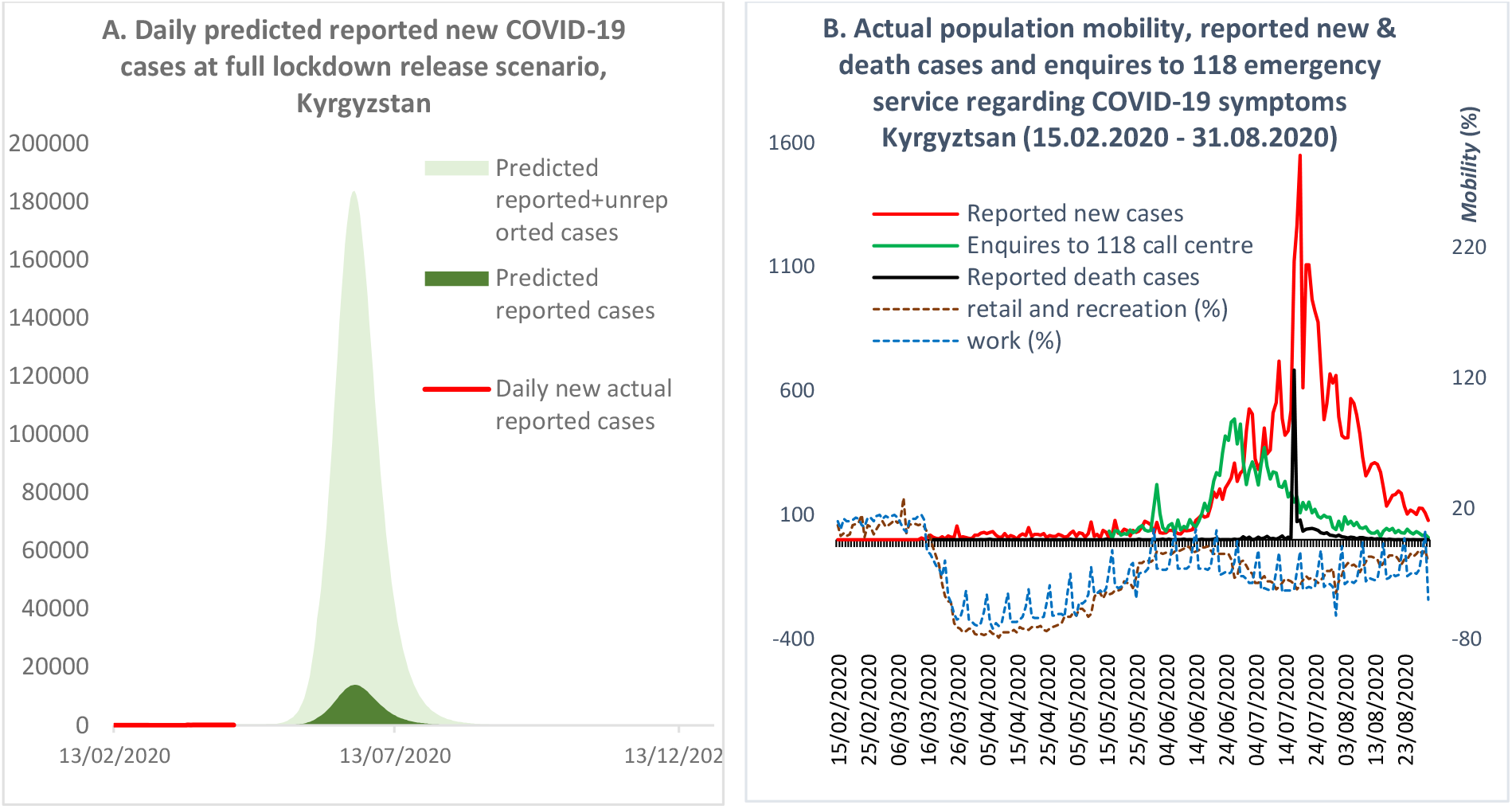
(A) Predicted and actual new cases after releasing the lockdown. (B) Actual new and death cases after releasing the lockdown and population mobility. Resources for Figure 2B: Kyrgyzstan MoH [1]; Google mobility report [10].

The actual course of the epidemic after the release of lockdown took a similar pattern to that predicted by the model. As shown in Figure 2B, with the lifting of lockdown, population mobility began returning to the normal mode, and by the beginning of June mobility had reached the pre-lockdown level. With the intensification of the population’s mobility, the number of reported new cases and deaths began to increase, reaching a peak in the middle of July. It should be noted that until July 17, 2020, the official statistics of new cases included PCR-positive tests only, although the number of cases diagnosed with pneumonia were exceeding the PCR confirmed cases. By the end of July, the proportion of cases with PCR-positive tests comprised just 12%; the remainder were diagnosed with pneumonia [11]. On July 17, 2020, the MoH officially recognised patients with pneumonia as COVID-19 cases and combined the two statistics [12].

A similar pattern to the reported cases was observed with enquires to the 118 ‘hotline’ about COVID-19 symptoms, although the peak of calls occurred in the middle of June (Figure 2B). It is important to note that the 118 hotline was one of many state and private emergency call centres, which were not included in our analysis due to difficulties accessing their data.

Based on the above, we consider that the model accurately predicted the timing of the actual epidemic peak following the lifting of the lockdown in May. However, the magnitude of the epidemic peak predicted by the model was observed to be larger than the actual occurrence. As shown in Figure 2A, the model predicted that reported new cases would reach 14,000 per day during the peak, which was about ten-times higher than the actual officially reported statistics.

### How accurately did the model predict hospital demand and occupancy and their effect on mortality for the full lockdown release scenario?

With the existing hospital capacity [13] and full lockdown release in May, the simulation predicted that the health system would become overwhelmed due to an extensive influx of patients during the first part of July. According to Figure 3 (A, B and C, respectively) the predicted demand for ICU beds, ventilators, and surge beds would far exceed their availability. For example, the daily demand for surge beds would reach 20,000, whereas the number of surge beds available was 2,200 [13]. At the same time, the occupancy of surge beds would be higher than the demand, as this included the available beds, occupied by those who required ICU and ventilation/oxygen treatment but could not access them, as well as the additional beds created in general wards (e.g. additional beds in corridors, or in additional temporary hospitals). In contrast to the surge bed situation, the occupancy of ICU beds and ventilators would barely exceed their availability thresholds due to the reduced flexibility for creating additional spaces in ICU wards and obtaining additional ventilators.

**Figure 3.**
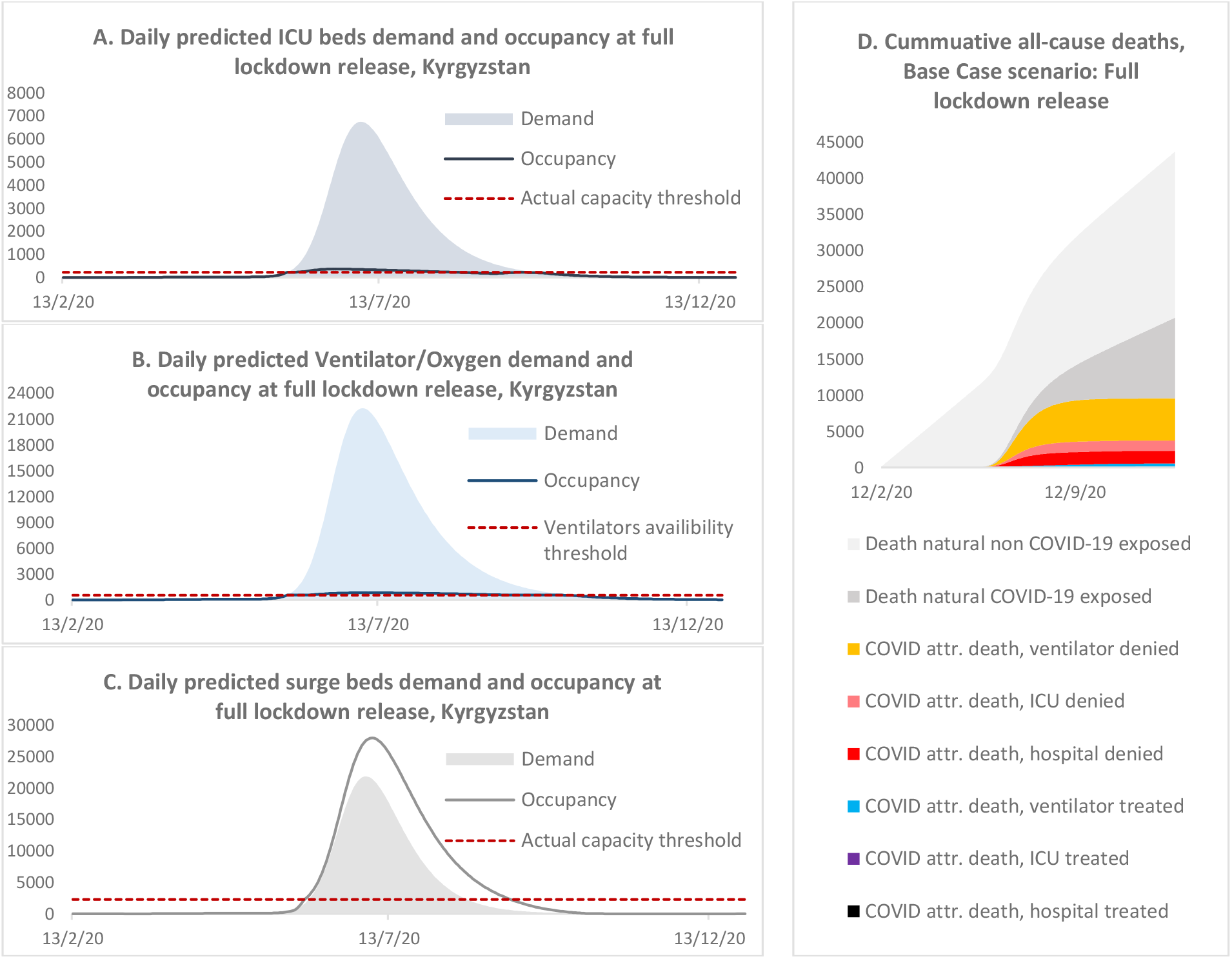
Daily predicted ICU bed (A), ventilator/oxygen (B), and surge bed (C) demand and occupancy and (D) cumulative deaths for the full lockdown release scenario.

With the increased pressure on the existing health system, the model predicted an increase in deaths, which in the baseline scenario would reach 6300 by the end of the simulation period. In Figure 3(D), we stratified the expected cumulative mortality into several categories, to reflect the contribution of deaths attributable to COVID-19 to all-cause mortality and to analyse the level of mortality among patients with severe COVID-19 symptoms who received treatment compared with those who could not access the required treatment and resources. The model predicted that the highest proportion of mortality would be among those individuals who did not have access to oxygen treatment, followed by denial of ICU and surge beds. According to the model, the potential contribution of mortality attributable to COVID-19 to all-cause mortality was not as high as in countries with a larger proportion of older people. In the baseline scenario, deaths attributable to COVID-19 would increase the yearly all-cause mortality statistics by about 22.7%. However, it is important to be aware that the model projections did not account for any interplay between COVID-19 with other diseases or factors; therefore, deaths not attributable to COVID-19 were assumed not to be affected by the COVID-19 epidemic.

The actual situation with the health system echoed the predicted full lockdown release scenario. The rapid increase in symptomatic cases put tremendous pressure on the health system, which was not prepared for such a heavy influx of patients with severe symptoms, most of whom required treatment with oxygen [11,14,15]. As predicted by the model, the peak of hospital occupancy occurred in July, with a difference of about two weeks compared with the model’s prediction, and comprised 19,774 patients per day as of July 18, 2020 [14].

In addition, hospitals experienced acute shortages of medical staff, personal protective equipment (PPE) and medicaments. Many of the specialists who were available became infected with COVID-19 during the course of their work. Thus, on August 3, 2020, the MoH reported that medical workers comprised approximately 16.8% of all COVID-19 cases, of whom about 43.7% were nurses or lab technicians and 34.7% were doctors [16]. As a result, many patients could not access hospital treatment or medical resources, which contributed to the increase in the number of deaths. The official statistics reported that there were 1362 deaths during June and July [14]. Available evidence suggests that COVID-19 was estimated to be the third most common cause of death in Kyrgyzstan during 2020, with an average of 22 deaths per 100,000 population nationally and 59 cases per 100,000 population in the capital city, Bishkek, which suffered the most during the epidemic [17]. Most of the deaths attributable to COVID-19 were among individuals with severe symptoms of pneumonia. According to the National Statistical Committee, by the time of the peak of the epidemic in July, deaths from pneumonia exceeded the average annual levels seen in previous years. There were 598, 646 and 626 deaths from pneumonia per year in 2017, 2018 and 2019, respectively, whereas between 1 and 17 July, 2020, there were 610 deaths from pneumonia and a total of 887 deaths from the beginning of the year to July 17 [12].

Based on the above, we consider that the model could predict the approximate timing of increased pressure on the health system, as well as insufficiencies in the number of available surge and ICU beds and ventilators/oxygen equipment during the peak of the epidemic. However, the model predicted a higher magnitude of demand for hospital treatment and occupancy than the officially reported actual situation. Moreover, the model’s flexibility for predicting occupancy for ICU and oxygen treatment was lower compared with the actual occurrence, when the lack of surge beds and oxygen devices was improved by the mobilisation of funds from the public and private sectors.

## Discussion

The assessment of the hypothetical full lockdown release scenario against the actual course of the epidemic following the release of the strict measures on May 10, 2020, showed that the timing of the model output prediction of the peak had a difference of two weeks compared with the timing of the actual occurrence of the peak. The model’s estimation of the magnitude of the epidemic peak was a few times higher compared with what actually happened during the outbreak. However, it is important to be aware that the official reports of case numbers remain debatable. Hence, some experts estimate the actual number of new symptomatic cases to be at least ten times higher than that of the official reports [18]. The Republican Scientific Centre on Infection Control, under the MoH, estimated there were 1,860,000 cases by the end of June, 2020 [18]. Considering that almost 20% of cases may develop symptoms, the number of symptomatic cases could have reached 370,000, whereas the official number of new cases was 32,000 by the end of July, which included only those individuals who tested positive by PCR or showed symptoms of pneumonia [14].

According to the model, an extensive increase in new cases would result in the health system being overwhelmed and, as a consequence, high mortality rates, which were also overestimated compared with the official data [18]. However, some experts consider that, as with the number of reported new cases, the actual magnitude of deaths was much higher than the reported statistics. According to some health specialists, during the peak of the epidemic in July, 2020, many people may have died at home and those cases were therefore not included in the official statistics [19].

Finally, the situation with the health system during the peak of the epidemic echoed the outcomes predicted by the model, with many patients unable to access hospital treatment and resources as a result of the acute shortages in surge and ICU beds and oxygen aid facilities and equipment, which also led to an increase in the number of deaths. The circumstances around the shortage of surge beds, oxygen equipment and medicaments have gradually improved as a result of tremendous support from the general population and the private sector, who managed to mobilise funds and resources and establish temporary ambulatory hospitals in hotels, sports centres and schools. As a result, in the capital city Bishkek alone, about 62,300 patients were able to receive ambulatory medical support by the end of July, 2020 [18].

Thus, based on the above, we can be confident that the model’s predictions could accurately reflect the actual timing of the epidemic curve, the magnitude of the epidemic peak and the pressure on the health system. However, there are some limitations of this model, associated with a number of uncertainties and assumptions about this novel disease and the effects of related interventions, that must be taken into account. One such limitation is that the model reflected the medium-term projection of the epidemic, where seasonality was not considered due to the limited evidence for this at the beginning of the year. Moreover, contrary to the model’s predictions, Kyrgyzstan experienced a second wave of the epidemic during October and November, as a result of nationwide protests and mass gatherings against the results of parliamentary elections. There were also some methodological limitations. The model was visually fitted, as part of the simulation process, through a web-based application. The particle filtering method has only recently became available, which we plan to apply for further simulations of the epidemic in Kyrgyzstan.

Accordingly, it remains unclear whether the visual fitting was appropriate for forecasting the epidemic. However, it is important to note that the primary function of the model was to support real-time decision-making, which urgently required evidence and tools to address the constantly changing situation with regards to the epidemic. Thus, in this use case, the model was fit for purpose, from a qualitative point of view, with its predictions matching the observed outcomes of the decision to release the lockdown in Kyrgyzstan.

## Data Availability

All relevant data are within the manuscript and its Supporting Information files

## Acknowledgement

We thank all who have collected, prepared, and shared data for this analysis. We are particularly thankful to our colleagues from the COVID-19 Modelling Consortium Dr. R.Aguas and Dr. O.Celhay for the computer programming of the model and developing the web-based application, Professor P.Ariana and Dr. R. Shretta for the help with preparing and critical review of the policy notes, and Dr. Adam Bodley for the scientific editing of the manuscript.

## References

1. Stop COVID KG. COVID-19 Kyrgyzstan [Internet]. 2020 [cited 2020 Jul 27]. Available from: https://covid.kg/

2. CoMo Consortium. The COVID-19 International Modelling Consortium [Internet]. 2020 [cited 2020 Dec 7]. Available from: https://como.bmj.com/

3. Aguas R, Hupert N, Shretta R, Celhay O, Moldokmatova A, Arifi F, et al. COVID-19 pandemic modelling in context: uniting people and technology across nations. BMJ Global Health (forthcoming).

4. KLOOP. «Бюрократическое алиби» и психологические ловушки: как Кыргызстан допустил вспышку коронавируса [Internet]. Kloop KG. 2020 [cited 2020 Nov 8]. Available from: https://kloop.kg/blog/2020/07/14/byurokraticheskoe-alibi-i-psihologicheskie-lovushki-kak-kyrgyzstan-dopustil-vspyshku-koronavirusa/

5. Kucharski AJ, Klepac P, Conlan AJK, Kissler SM, Tang ML, Fry H, et al. Effectiveness of 1. isolation, testing, contact tracing, and physical distancing on reducing transmission of SARS-CoV-2 in different settings: a mathematical modelling study. Lancet Infect Dis [Internet]. 2020 Oct 1 [cited 2020 Nov 10];20(10):1151–60. Available from: www.thelancet.com/infection

6. Day M. Covid-19: identifying and isolating asymptomatic people helped eliminate virus in Italian village. BMJ [Internet]. 2020 Mar 23 [cited 2020 Nov 10];368:m1165. Available from: http://group.bmj.com/group/rights-licensing/

7. Day M. Covid-19: four fifths of cases are asymptomatic, China figures indicate. BMJ [Internet]. 2020 Apr 2 [cited 2020 Aug 28];369:m1375. Available from: http://group.bmj.com/group/rights-licensing/

8. Oran DP, Topol EJ. Prevalence of Asymptomatic SARS-CoV-2 Infection?: A Narrative Review [Internet]. Vol. 173, Annals of internal medicine. NLM (Medline); 2020 [cited 2020 Nov 10]. p. 362–7. Available from: https://www.acpjournals.org/doi/abs/10.7326/M20-3012

9. Davies NG, Klepac P, Liu Y, Prem K, Jit M, Eggo RM. Age-dependent effects in the transmission and control of COVID-19 epidemics. Nat Med. 2020 Aug 16;26(8).

10. Google. COVID-19 Community Mobility Reports [Internet]. 2020 [cited 2020 Aug 6]. Available from: https://www.google.com/covid19/mobility/

11. Radio Azzattyk. В Кыргызстане с 1 октября резко изменилась ?;труктура эпидемии коронавируса [Internet]. Radio Azattyk. 2020 [cited 2020 Nov 11]. Available from: https://rus.azattyk.org/a/30914691.html

12. NSC KR. Mortality from pneumonia, 2019-2020 [Internet]. National Statistical 1. Committee of the Kyrgyz Republic . 2020 [cited 2020 Nov 8]. Available from: http://stat.kg

13. MoH KR. MoH Prikaz #181 on health system preparedness for the COVID-19 epidemic. March 23, 2020 [Internet]. 2020 [cited 2020 Aug 3]. Available from: http://med.kg/ru/dokumenty/prikazy.html

14. Akipress. Коронавирус в Кыргызстане. АКИpress [Internet]. Akipress. Kyrgyzstan . 2020 [cited 2020 Nov 13]. Available from: https://akipress.org/dolbor/covid-19/graph-kg/?hl=ru&from=covid-19&from=left

15. MoH KR. Ministry of Health of the Kyrgyz Republic / Министерство здравоохранения Кыргызской Республики [Internet]. 2020 [cited 2020 Aug 28]. Available from: http://med.kg/ru/

16. Kabar. В Кыргызстане 16,8 % заболевших коронавирусом – медики [Internet]. Kabar. 2020 [cited 2020 Nov 13]. Available from: http://kabar.kg/news/v-kyrgyzstane-16-8-zabolevshikh-koronavirusom-mediki/

17. Kaktus. Коронавирус - третья по масштабу причина смертности в Кыргызстане [Internet]. Kaktus KG. 2020 [cited 2020 Nov 13]. Available from: https://kaktus.media/doc/418321_koronavirys_tretia_po_masshtaby_prichina_smertn osti_v_kyrgyzstane.html

18. Radio Azattyk. Переболевших COVID-19 в Кыргызстане гораздо больше, чем приводится в официальной статистике? [Internet]. Radio Azattyk. 2020 [cited 2020 Nov 13]. Available from: https://rus.azattyk.org/a/30742521.html

19. Radio Azattyk. Статистика по COVID-19 снижается. Стоит ли доверять официальным данным? [Internet]. Radio Azattyk. 2020 [cited 2020 Nov 13]. Available from: https://rus.azattyk.org/a/30855114.html

20. Otto AM. COVID-19 update: Transmission 5% or less among close contacts [Internet]. The Hospitalist. 2020 [cited 2020 Aug 28]. Available from: https://www.the-hospitalist.org/hospitalist/article/218769/coronavirus-updates/covid-19-update-transmission-5-or-less-among-close

21. DRCU. Disaster Response Coordination Unit in the Kyrgyz Republic. Press Centre / Нацинальный штаб по противодействию эпидемии COVID-19 в Кыргызской Республике. Пресс центр [Internet]. 2020 [cited 2020 Aug 28]. Available from: https://www.gov.kg/ru/p/covid-19

22. Prem K, Cook AR, Jit M. Projecting social contact matrices in 152 countries using contact surveys and demographic data. PLoS Comput Biol [Internet]. 2017 Sep 1 [cited 2020 Aug 3];13(9):e1005697. Available from: http://www.dhsprogram.comhttp://data.worldbank.org/,UIS.Stathttp://data.uis.unesco.org,http://unstats.un.org/unsd/default.htm/,InternationalLaborOrganization http://www.ilo.org/global/statistics-and-databases/lang-en/index.htm.

23. NSTKG. National Statistical Committee of the Kyrgyz Republic [Internet]. 2020 [cited 2020 Aug 3]. Available from: http://www.stat.kg/en/

24. Linton N, Kobayashi T, Yang Y, Hayashi K, Akhmetzhanov A, Jung S, et al. Incubation Period and Other Epidemiological Characteristics of 2019 Novel Coronavirus Infections with Right Truncation: A Statistical Analysis of Publicly Available Case Data. J Clin Med [Internet]. 2020 Feb 17 [cited 2020 Aug 28];9(2):538. Available from: https:/pmc/articles/PMC7074197/?report=abstract

25. Khalili M, Karamouzian M, Nasiri N, Javadi S, Mirzazadeh A, Sharifi H. Epidemiological Characteristics of COVID-19: A Systemic Review and Meta-Analysis. medRxiv [Internet]. 2020 Apr 6 [cited 2020 Aug 28];2020.04.01.20050138. Available from: https://doi.org/10.1101/2020.04.01.20050138

26. Bi Q, Wu Y, Mei S, Ye C, Zou X, Zhang Z, et al. Epidemiology and Transmission of COVID-19 in Shenzhen China: Analysis of 391 cases and 1,286 of their close contacts. medRxiv [Internet]. 2020 Mar 27 [cited 2020 Aug 28];2020.03.03.20028423. Available from: https://doi.org/10.1101/2020.03.03.20028423

27. Mizumoto K, Kagaya K, Zarebski A, Chowell G. Estimating the Asymptomatic Proportion of 2019 Novel Coronavirus onboard the Princess Cruises Ship, 2020 [Internet]. medRxiv. Cold Spring Harbor Laboratory Press; 2020 Mar [cited 2020 Aug 28]. Available from: http://medrxiv.org/content/early/2020/03/06/2020.02.20.20025866.abstract

28. ECDC. Novel coronavirus disease 2019 (COVID-19) pandemic: increased transmission in the EU/EEA and the UK – sixth update. 2020.

29. Verity R, Okell LC, Dorigatti I, Winskill P, Whittaker C, Imai N, et al. Estimates of the severity of coronavirus disease 2019: a model-based analysis. Lancet Infect Dis [Internet]. 2020 Jun 1 [cited 2020 Aug 28];20(6):669–77. Available from: www.thelancet.com/infectionVolwww.thelancet.com/infectionVol

30. Richardson S, Hirsch JS, Narasimhan M, Crawford JM, McGinn T, Davidson KW, et al. Presenting Characteristics, Comorbidities, and Outcomes among 5700 Patients Hospitalized with COVID-19 in the New York City Area. JAMA - J Am Med Assoc [Internet]. 2020 May 26 [cited 2020 Aug 28];323(20):2052–9. Available from: /pmc/articles/PMC7177629/?report=abstract

31. Petrilli CM, Jones SA, Yang J, Rajagopalan H, O’Donnell LF, Chernyak Y, et al. Factors associated with hospitalization and critical illness among 4,103 patients with COVID-19 disease in New York City. medRxiv [Internet]. 2020 Apr 11 [cited 2020 Aug 28];2020.04.08.20057794. Available from: https://doi.org/10.1101/2020.04.08.20057794

32. Zhou F, Yu T, Du R, Fan G, Liu Y, Liu Z, et al. Clinical course and risk factors for mortality of adult inpatients with COVID-19 in Wuhan, China: a retrospective cohort study. Lancet [Internet]. 2020 Mar 28 [cited 2020 Aug 28];395(10229):1054–62. Available from: https://doi.org/10.1016/

33. Wang Z, Ji J, Liu Y, Liu R, Zha Y, Chang X, et al. Survival analysis of hospital length of stay of novel coronavirus (COVID-19) pneumonia patients in Sichuan, China. medRxiv [Internet]. 2020 Apr 10 [cited 2020 Aug 28];2020.04.07.20057299. Available from: https://doi.org/10.1101/2020.04.07.20057299

34. UN. World Population Prospects: Population Division [Internet]. 2019 [cited 2020 Aug 3]. Available from: https://population.un.org/wpp/Download/Standard/Population/

35. RPS. Handwashing [Internet]. Royal Pharmaceutical Society of Great Britain. 2020 [cited 2020 Aug 28]. Available from: https://www.rpharms.com/resources/pharmacy-guides/ams-portal/handwashing

