## Supplementary material for "Mathematical modelling projections versus the actual course of the COVID-19 epidemic following the nationwide lockdown in Kyrgyzstan": S2 Appendix

**S2 Appendix. Parameters and visual fitting charts**

**Part I. Model base indicators**

A. **General**

| **Description** | **Value** | **Resource** |
| --- | --- | --- |
| Probability of infection given contact: (0 to 0.2) | 0.033 | Visually calibrated and based on: [20] |
| Percentage of all asymptomatic infections that are reported: | 0 | Assumed based on: [1] |
| Percentage of all symptomatic infections that are reported: | 12 | Visually calibrated and based on: [1,13,21] |
| Percentage of all hospitalisations that are reported: | 90 | Assumed based on: [1,21] |
| Social contacts data (country): | Kyrgyzstan | [22] |
| Mean household size: | 4.2 | [23] |
| Mean number of infectious migrants per day: | 10^{-5} | Assumed based on: [21] |

**Figure S2.** Visual fitting of the projected epidemic curve against actual reported cumulative deaths (A) and daily new cases of COVID-19 (B) as of April 24, 2020.

A) **B)**

Predicted vs.actual cumulative cases

Cummulative death cases

|  | Predicted Cummulative death cases | Actual cumulative death cases |
| --- | --- | --- |
| 2020-02-13 | 0 |  |
| 2020-02-14 | 0 |  |
| 2020-02-15 | 0 |  |
| 2020-02-16 | 0 |  |
| 2020-02-17 | 0 |  |
| 2020-02-18 | 0 |  |
| 2020-02-19 | 0 |  |
| 2020-02-20 | 0 |  |
| 2020-02-21 | 0 |  |
| 2020-02-22 | 0 |  |
| 2020-02-23 | 0 |  |
| 2020-02-24 | 0 |  |
| 2020-02-25 | 0 |  |
| 2020-02-26 | 0 |  |
| 2020-02-27 | 0 |  |
| 2020-02-28 | 0 |  |
| 2020-02-29 | 0 |  |
| 2020-03-01 | 0 |  |
| 2020-03-02 | 0 |  |
| 2020-03-03 | 0 |  |
| 2020-03-04 | 0 |  |
| 2020-03-05 | 0 |  |
| 2020-03-06 | 0 |  |
| 2020-03-07 | 0 |  |
| 2020-03-08 | 0 |  |
| 2020-03-09 | 0 |  |
| 2020-03-10 | 0 |  |
| 2020-03-11 | 0 |  |
| 2020-03-12 | 0 |  |
| 2020-03-13 | 0 |  |
| 2020-03-14 | 0 |  |
| 2020-03-15 | 0 |  |
| 2020-03-16 | 0 |  |
| 2020-03-17 | 0 |  |
| 2020-03-18 | 0 |  |
| 2020-03-19 | 0 |  |
| 2020-03-20 | 0 |  |
| 2020-03-21 | 0 |  |
| 2020-03-22 | 0 |  |
| 2020-03-23 | 0 |  |
| 2020-03-24 | 0 |  |
| 2020-03-25 | 0 |  |
| 2020-03-26 | 1 |  |
| 2020-03-27 | 1 |  |
| 2020-03-28 | 1 |  |
| 2020-03-29 | 1 |  |
| 2020-03-30 | 1 |  |
| 2020-03-31 | 1 |  |
| 2020-04-01 | 1 |  |
| 2020-04-02 | 2 |  |
| 2020-04-03 | 2 |  |
| 2020-04-04 | 2 | 1 |
| 2020-04-05 | 2 | 2 |
| 2020-04-06 | 3 | 3 |
| 2020-04-07 | 3 |  |
| 2020-04-08 | 3 | 5 |
| 2020-04-09 | 4 |  |
| 2020-04-10 | 4 |  |
| 2020-04-11 | 4 |  |
| 2020-04-12 | 5 | 6 |
| 2020-04-13 | 5 |  |
| 2020-04-14 | 5 |  |
| 2020-04-15 | 6 |  |
| 2020-04-16 | 6 |  |
| 2020-04-17 | 7 |  |
| 2020-04-18 | 7 |  |
| 2020-04-19 | 7 |  |
| 2020-04-20 | 8 |  |
| 2020-04-21 | 8 |  |
| 2020-04-22 | 9 | 8 |
| 2020-04-23 | 9 |  |
| 2020-04-24 | 9 |  |
| 2020-04-25 | 10 | 10 |
| 2020-04-26 | 10 |  |
| 2020-04-27 | 11 |  |
| 2020-04-28 | 11 |  |
| 2020-04-29 | 12 |  |

Predicted vs.actual daily new cases

Daily new cases

|  | Predicted daily reported new cases | Predicted daily reported+unreported new cases | Actual daily reported new cases |
| --- | --- | --- | --- |
| 2020-02-13 | 0 | 0 |  |
| 2020-02-14 | 0 | 0 |  |
| 2020-02-15 | 0 | 0 |  |
| 2020-02-16 | 0 | 0 |  |
| 2020-02-17 | 0 | 0 |  |
| 2020-02-18 | 0 | 0 |  |
| 2020-02-19 | 0 | 0 |  |
| 2020-02-20 | 0 | 1 |  |
| 2020-02-21 | 0 | 1 |  |
| 2020-02-22 | 0 | 1 |  |
| 2020-02-23 | 0 | 1 |  |
| 2020-02-24 | 0 | 1 |  |
| 2020-02-25 | 0 | 2 |  |
| 2020-02-26 | 0 | 2 |  |
| 2020-02-27 | 0 | 2 |  |
| 2020-02-28 | 0 | 3 |  |
| 2020-02-29 | 0 | 3 |  |
| 2020-03-01 | 0 | 4 |  |
| 2020-03-02 | 0 | 5 |  |
| 2020-03-03 | 1 | 6 |  |
| 2020-03-04 | 1 | 8 |  |
| 2020-03-05 | 1 | 9 |  |
| 2020-03-06 | 1 | 11 |  |
| 2020-03-07 | 1 | 14 |  |
| 2020-03-08 | 1 | 17 |  |
| 2020-03-09 | 2 | 21 |  |
| 2020-03-10 | 2 | 26 |  |
| 2020-03-11 | 3 | 32 |  |
| 2020-03-12 | 3 | 39 |  |
| 2020-03-13 | 4 | 48 |  |
| 2020-03-14 | 5 | 60 | 7 |
| 2020-03-15 | 6 | 73 | 2 |
| 2020-03-16 | 7 | 90 | 7 |
| 2020-03-17 | 9 | 111 | 19 |
| 2020-03-18 | 11 | 137 | 11 |
| 2020-03-19 | 14 | 169 | 5 |
| 2020-03-20 | 17 | 208 | 4 |
| 2020-03-21 | 21 | 257 | 12 |
| 2020-03-22 | 26 | 316 | 4 |
| 2020-03-23 | 32 | 390 | 15 |
| 2020-03-24 | 39 | 480 | 10 |
| 2020-03-25 | 48 | 592 | 11 |
| 2020-03-26 | 46 | 557 | 55 |
| 2020-03-27 | 45 | 537 | 9 |
| 2020-03-28 | 45 | 526 | 12 |
| 2020-03-29 | 45 | 520 | 8 |
| 2020-03-30 | 45 | 517 | 5 |
| 2020-03-31 | 45 | 517 | 5 |
| 2020-04-01 | 46 | 526 | 19 |
| 2020-04-02 | 45 | 513 | 19 |
| 2020-04-03 | 44 | 502 | 26 |
| 2020-04-04 | 44 | 494 | 30 |
| 2020-04-05 | 43 | 486 | 25 |
| 2020-04-06 | 43 | 480 | 30 |
| 2020-04-07 | 42 | 473 | 32 |
| 2020-04-08 | 41 | 467 | 19 |
| 2020-04-09 | 41 | 461 | 10 |
| 2020-04-10 | 40 | 455 | 11 |
| 2020-04-11 | 40 | 450 | 24 |
| 2020-04-12 | 39 | 444 | 9 |
| 2020-04-13 | 39 | 438 | 16 |
| 2020-04-14 | 38 | 432 | 12 |
| 2020-04-15 | 38 | 427 | 2 |
| 2020-04-16 | 37 | 421 | 17 |
| 2020-04-17 | 37 | 416 | 23 |

B. **Disease parameters**

| **Description** | **Value** | **Resource** |
| --- | --- | --- |
| Average incubation period: (1 to 7 days) | 3.5 | [24–26] |
| Average duration of symptomatic infection period: (1 to 7 days) | 4.5 | [24] |
| Month of peak infectivity of the virus: (1, 2, …, 12) | 0 | NA (disabled parameter) |
| Annual variation in infectivity of the virus: | 0 | NA (disabled parameter) |
| Average duration of immunity: (0.5 to 150) | 150 | NA (disabled parameter) |
| Probability upon infection of developing clinical symptoms: | 0.55 | [7,27,28] |
| Probability upon hospitalisation of requiring ICU admission: | 0.50 | [29–31] |
| Probability upon admission to the ICU of requiring a ventilator: | 0.75 | [30] |

C. **Hospitalisation**

| **Description** | **Value** | **Unit** | **Resource** |
| --- | --- | --- | --- |
| Maximum number of hospital surge beds | 2200 | beds | [13] |
| Maximum number of ICU beds without ventilators | 226 | beds | [13] |
| Maximum number of ventilators | 551 | ventilators | [13] |
| Relative percentage of regular daily contacts when hospitalised: | 15 | % | Estimated in consultation with local clinical hospitals |
| Scaling factor for infection hospitalisation rate: (0.5 to 4) | 1 |  | NA (disabled parameter) |
| Probability of dying when hospitalised (not requiring oxygen): | 5 | % | Estimated in consultation with MoH, local clinical hospitals |
| Probability of dying when hospitalised if requiring oxygen: | 15 | % | Estimated in consultation with MoH, local clinical hospitals |
| Probability of dying when denied hospitalisation (not requiring oxygen): | 20 | % | Estimated in consultation with MoH, local clinical hospitals |
| Probability of dying when denied hospitalisation if requiring oxygen: | 40 | % | Estimated in consultation with MoH, local clinical hospitals |
| Probability of dying when admitted to ICU (not requiring oxygen): | 30 | % | Estimated in consultation with MoH, local clinical hospitals and based on: (31) |
| Probability of dying when admitted to ICU if requiring oxygen: | 55 | % | Estimated in consultation with MoH, local clinical hospitals and based on: (31) |
| Probability of dying when admission to ICU denied (not requiring oxygen): | 70 | % | Estimated in consultation with MoH, local clinical hospitals |
| Probability of dying when admission to ICU denied if requiring oxygen: | 75 | % | Estimated in consultation with MoH, local clinical hospitals |
| Probability of dying when ventilated: | 70 | % | Estimated in consultation with MoH, local clinical hospitals and based on: (31) |
| Probability of dying when ventilator denied: | 95 | % | Estimated in consultation with MoH, local clinical hospitals |
| Duration of hospitalised infection: (1 to 30) | 14 | days | Estimated in consultation with MoH, local clinical hospitals and based on: [24,25,32,33] |
| Duration of ICU infection: (1 to 30) | 20 | days | Estimated in consultation with MoH, local clinical hospitals and based on: [24,25,32,33] |
| Duration of ventilated infection: (1 to 30) | 24 | days | Estimated in consultation with MoH, local clinical hospitals and based on: [24,25,32,33] |

D. **Age-based fatality rate and infections that lead to hospitalisation**

| Age category/resource (years) | Age-based relative fatality rate in a well-resourced scenario (%) | Age-stratum-specific hospitalisation (proportion of all (asymptomatic + symptomatic) infections that lead to hospitalisation) (%) |
| --- | --- | --- |
|  | [29] | [29] |
| 0–4 | 0.0016 | 0 |
| 5–9 | 0.0016 | 0 |
| 10–14 | 0.007 | 0.04 |
| 15–19 | 0.007 | 0.04 |
| 20–24 | 0.031 | 1.1 |
| 25–29 | 0.031 | 1.1 |
| 30–34 | 0.26 | 3.43 |
| 35–39 | 0.26 | 3.43 |
| 40–44 | 0.48 | 4.25 |
| 45–49 | 0.48 | 4.25 |
| 50–54 | 0.6 | 8.2 |
| 55–59 | 0.6 | 8.2 |
| 60–64 | 1.9 | 11.8 |
| 65–69 | 1.9 | 11.8 |
| 70–74 | 4.3 | 16.6 |
| 75–79 | 4.3 | 16.6 |
| 80–84 | 7.8 | 18.4 |
| 85–89 | 7.8 | 18.4 |
| 90–94 | 7.8 | 18.4 |
| 95–99 | 7.8 | 18.4 |
| 100+ | 7.8 | 18.4 |

E. **Population structure, birth and death rates**

| Age category | Population | Number of births per person (i.e. 0.5* births per woman) per day | Deaths per person per day |
| --- | --- | --- | --- |
|  | [34] | [34] | [34] |
| 0–4 | 760,255 | 0.0 | 0.0000101605 |
| 5–9 | 769,195 | 0.0 | 0.0000007304 |
| 10–14 | 600,626 | 0.0 | 0.0000009053 |
| 15–19 | 500,075 | 0.0000448688 | 0.0000015779 |
| 20–24 | 514,389 | 0.0002696000 | 0.0000022355 |
| 25–29 | 568,551 | 0.0002296603 | 0.0000027381 |
| 30–34 | 572,187 | 0.0001459984 | 0.0000038920 |
| 35–39 | 442,518 | 0.0000888265 | 0.0000059593 |
| 40–44 | 361,459 | 0.0000250881 | 0.0000095105 |
| 45–49 | 324,976 | 0.0000012351 | 0.0000134190 |
| 50–54 | 295,973 | 0.0 | 0.0000201361 |
| 55–59 | 285,462 | 0.0 | 0.0000270330 |
| 60–64 | 220,069 | 0.0 | 0.0000408832 |
| 65–69 | 143,755 | 0.0 | 0.0000581985 |
| 70–74 | 78,619 | 0.0 | 0.0001209308 |
| 75–79 | 32,595 | 0.0 | 0.0004064232 |
| 80–84 | 32,507 | 0.0 | 0.0004353677 |
| 85–89 | 15,592 | 0.0 | 0.0004875172 |
| 90–94 | 4,435 | 0.0 | 0.0005742387 |
| 95–99 | 897 | 0.0 | 0.0006177715 |
| 100+ | 56 | 0.0 | 0.0098953750 |

**Part II. Intervention indicators**

A. **Intervention adherence/efficacy**

| **Category** | **Description** | **Value** | **Resource** |
| --- | --- | --- | --- |
| Self-isolation if symptomatic | Adherence: | 50 | Assumed following consultations with local experts |
| Screening (index tracing) | Overdispersion: (1, 2, 3, 4 or 5) | 4 | Assumed following consultations with local experts and based on [13] |
|  | Test sensitivity: | 80 | Based on: [13] and consultations with local experts |
| Household isolation (when symptomatic) | Days in isolation for an average person: | 14 | Assumed following consultations with local experts and based on: [21] |
|  | Days to implement maximum quarantine coverage: (1 to 5) | 2 | Assumed following consultations with local experts |
|  | Decrease in the number of other contacts when quarantined: | 10 | Assumed following consultations with local experts |
|  | Increase in the number of contacts at home when quarantined: | 100 | Assumed following consultations with local experts |
| Social distancing | Adherence: | 100 | Based on the assumption that all following social distancing will have been doing it with 100% efficacy. Therefore, in hypothetical scenarios, this parameter will only count for such people |
| Handwashing | Efficacy: (0–35%) | 5 | Assumed following consultations with local experts and based on [35] |
| Mask wearing | Efficacy: (0–25%) | 5 | Assumed following consultations with local experts |
| Working at home | Efficacy: | 80 | Assumed following consultations with local experts |
|  | Home contact inflation due to working from home: | 10 |  |
| School closures | Efficacy: | 80 | Assumed following consultations with local experts |
|  | Home contact inflation due to school closure: | 10 |  |
| Shielding the elderly | Efficacy: | 95 | Assumed following consultations with local experts |
|  | Minimum age for shielding the elderly: (0 to 100) | 70 |  |
| International travel ban | Efficacy: | 80 | Assumed following consultations with local experts and based on: [21] |
