## Supplementary material for "Mathematical modelling projections versus the actual course of the COVID-19 epidemic following the nationwide lockdown in Kyrgyzstan": S1 Appendix

### Modelling the potential impact of various interventions on the COVID-19 epidemic in the Kyrgyz Republic

#### Policy Notes

A. Moldokmatova, A. Dooronbekova, Ch. Zhumalieva, A. Estebesova, A. Mukambetov, A. Kubatova, Sh. Ibragimov, T.Abdyldaev, P. Ariana, R. Aguas, R. Shretta, L. White

April 2020

### Executive Summary

Despite the set of measures introduced in the Kyrgyz Republic in response to COVID-19, including lockdown and closure of borders, the epidemic has spread across all regions, except Talas Oblast, with an alarming rate of infection among healthcare workers. There is an urgent need for a tailored, evidence-based strategy to inform decisions on effective response measures to COVID-19 for Kyrgyzstan. This brief presents preliminary findings of mathematical models used to project the course of the COVID-19 epidemic in the Kyrgyz Republic given various interventions.

The simulation is based on local epidemiological data as of **24 April 2020** and assumptions about current interventions, with an appreciation of local social contexts, as well as existing global evidence regarding the nature of the disease and its spread. There remain **many uncertainties** as evidence is rapidly being generated; thus, results will change as we learn more about the nature of the disease and the impact of interventions on disease outcomes and as we receive more reliable data about intervention intensity and coverage.

### Table of Contents

**Situation** 4

**Alternative interventions/scenarios** 5

**Limitations** 7

**Assumptions** 7

**Projected model outcomes** 8

**Acknowledgements** 14

**References** 15

### Situation

The number of cases of COVID-19 continues to increase in the Kyrgyz Republic. The first three cases were reported among travellers returning from a pilgrimage to Saudi Arabia on 16 March 2020. By 24 April, the number of cases had reached 729 (MoH, 2020a). The majority of cases (393) are concentrated in the south of the country (Osh and Jalalabad provinces), followed by Bishkek (180) and the remote Naryn province (94).

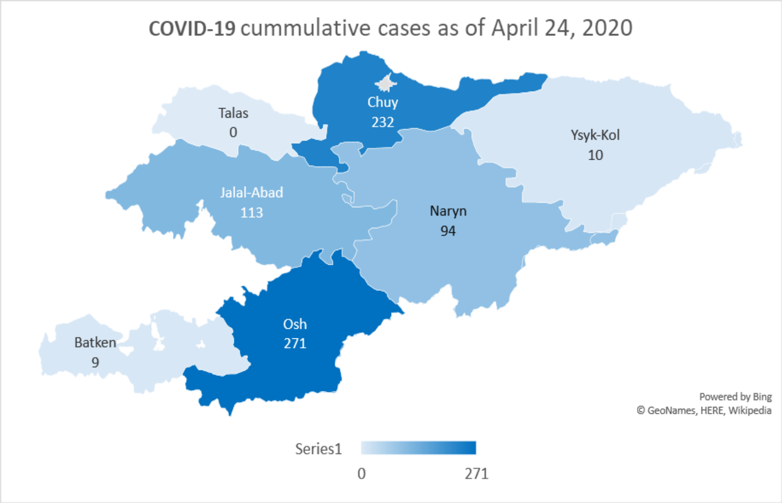


Unlike international trends, 71% of cases have occurred among those aged 20 to 60 years, while cases among those aged more than 60 years accounted for just 12% of cases. The distribution between the sexes is almost equal, with slightly higher rates among females (54%). Health workers account for 26% of cases. As of April 24, 2020, the Ministry of Health of Kyrgyz Republic (MoH KR) has reported eight deaths, mainly among elderly people aged more than 65 years or individuals with pre-existing health conditions. Thus, the case fatality rate (CFR) is 1.1% (MoH, 2020a).

The available data points to a decrease in imported cases but an increase in local transmission, despite the current lockdown and quarantine measures. More evidence is needed to determine the effectiveness of various interventions in the context of Kyrgyzstan. It is possible that the increased detection of cases may be due to improved diagnostic capacity and availability of testing resources.

According to the World Health Organization, different countries exhibit variations in the rates of disease severity, mortality and hospital admissions. In China, about 15% to 20% of cases required hospitalization, of whom 15% had severe symptoms and 5% required ventilation and other intensive care manipulations. In Italy and Spain, between 40% and 55% of positive cases were admitted to hospital, with between 7% and 12% needing intensive care (WHO, 2020b). These variations may be driven by factors such as population structure, efficiency of prevention and control measures, and preparedness and capacity of health systems.

MoH KR has developed a plan for the preparation of hospital capacity and reorganised existing hospitals to treat COVID-19 patients. In total, more than two thousand hospital beds, including 226 in intensive care units (ICUs) will be set up, in several stages (MOH Order #181, March 23, 2020). The current stock of respiratory ventilators is 625 devices, 74 of which require maintenance (MoH, 2020b).

The government responded rapidly to the COVID-19 pandemic and introduced emergency measures in two major cities (Bishkek and Osh) and in the affected Osh and Jalalabad provinces on 22 March, 2020, with a closure of borders; a travel ban; testing, isolation and quarantining; physical distancing; and health communication. In response to the growing rates of infection, the country declared a state of emergency from 25 March to mid-April and later extended the lockdown to the beginning of May. Stricter measures, including curfews, checkpoints and the closure of all businesses except essential ones (e.g. grocery stores, pharmacies and gas stations), were rolled-out under the state of emergency. As lockdowns can affect people's wellbeing, and socio-economic challenges increase, there is an urgent need for clear evidence to inform the country's next steps to tackle the pandemic, while acknowledging the wider health, social and economic consequences of any steps taken.

#### Alternative interventions/scenarios

In response to the current situation, the Kyrgyz modelling group, in cooperation with the international COVID-19 Modelling Consortium (CoMo Consortium) and the Soros Foundation in the Kyrgyz Republic, projected possible courses of the pandemic in the country, through modelling several scenarios with varying interventions. The team applied the mathematical modelling framework developed by the Oxford Modelling Group for Global Health (OMGH) in collaboration with the CoMo Consortium. The model can be used to estimate the impact of potential intervention strategies on the course of COVID-19 epidemics in individual countries and help to inform policy decisions. We included three levels of potential disruption to the social and economic situation in Kyrgyzstan (Table 1) and projected five scenarios with various interventions and timelines, to address the following questions:

- *What would be the scale of the epidemic if lockdown is fully lifted after 10 May, 2020?*
- *What would be the impact of 'low disruptive interventions' after lifting the lockdown on 10 May, 2020?*
- *What would be the impact of 'medium disruption interventions' after lifting the lockdown on 10 May, 2020?*
- *What would be the impact of 'high disruptive interventions' after lifting the lockdown on 10 May, 2020?*

***Table 1.*** *Level of disruption of intervention scenarios on social life and the economy.*

| Level of disruption | Scenario | Intervention |
|---|---|---|
| ***Low*** | Scenario 1 (Baseline) | Baseline: Full release |
| | Scenario 2 | Managed lower intensity release |
| ***Medium*** | Scenario 3 | Managed higher intensity release |
| ***High*** | Scenario 4 | Prolonged lockdown with full release |
| | Scenario 5 | Prolonged lockdown with managed release |

***Table 2***. *Intervention parameters for modelled hypothetical scenarios.*

| **Intervention** | **Hypothetical scenario** | | | | |
|---|---|---|---|---|---|
| | 1 | 2 | 3 | 4 | 5 |
| ***Initial full lockdown*** | 8 weeks | 8 weeks | 8 weeks | | |
| ***Extended full lockdown*** | | | | 12 weeks | |
| ***Extended full lockdown*** | | | | | 16 weeks |
| ***Additional post-lockdown measures*** | | | | | |
| ***Mask wearing*** *(coverage)* | 20% until the end of the simulation period | | | | |
| ***Hand washing*** *(coverage)* | 60% until the end of the simulation period | | | | |
| ***Self-isolation if symptomatic*** *(coverage)* | | 40% for 12 weeks | 60% for 16 weeks | | 60% for 23 weeks |
| ***Case tracing*** *(number of contacts per index case)* | | 20 for 12 weeks | 20 for 16 weeks | | 40 for 23 weeks |
| ***Household isolation if symptomatic*** *(coverage)* | | 30% for 12 weeks | 30% for 16 weeks | | 40% for 23 weeks |
| ***Social distancing*** *(coverage)* | | | 30% for 16 weeks | | 40% for 23 weeks |
| ***Working from home*** *(coverage)* | | | | | 30% for 14 weeks |
| ***School closure*** *(coverage)* | Summer holidays for 12 weeks (100%) | Summer holidays for 12 weeks (100%) | Summer holidays for 12 weeks (100%) | Summer holidays for 12 weeks (100%) | Summer holidays for 12 weeks (100%) + 80% for 6 weeks in new academic year |
| ***International travel ban*** *(coverage)* | | | | | 50% for 10 weeks |

#### Limitations

- We need to take into account uncertainties about the virus and its epidemiology, as well as assumptions regarding current intervention coverage and efficacy affected by social, cultural and economic factors. The model outputs will change as we learn more about the disease and the impact of interventions on the nature of the disease and as we receive more reliable data on intervention intensity and coverage.
- The current model did not include vaccination as a pharmaceutical intervention for the prevention of COVID-19 infection, although this option was foreseen and the modelling tool includes the assumption of its availability at a later date.
- We have not included seasonality due to a lack of evidence on whether this virus will exhibit a seasonal pattern and, if it does, whether this will be a similar pattern to that seen with influenza.
- Another important limitation of this projection is that it did not include an analysis of the effect of high rates of COVID-19 infection among healthcare workers on the health system's capacity to respond to the epidemic.

#### Assumptions

- The model is based on the epidemiological data available as of April 24, 2020 (MoH, 2020a). There is a need to continually update the simulations with new data/evidence.
- Due to the unavailability of direct values for intervention coverage, adherence and efficacy, the related model assumptions were based on other existing proxy data and information, including Google Maps analysis of community mobility in countries (Google Map, 2020), the EpiCOVID online survey in Central Asia (EpiCOVID, 2020) and weekly reports of the Disaster Response Coordination Unit in the Kyrgyz Republic (DRCU, 2020).
- The currently accepted global evidence on the nature of COVID-19 disease, which is yet to be updated, was used for the disease parameters (CDC China, 2020; Korean Society of Infectious Diseases et al., 2020; Liu et al., 2020; Riou et al., 2020; WHO, 2020a).
- The demographics parameter values for the population age structure were based on United Nations (UN) data for 2019 (UN, 2020).
- The social contact matrices projection for 152 countries (Prem et al., 2017) was used to estimate the contact patterns between different age groups in Kyrgyzstan.

Thus, the following outputs should be interpreted in light of the above assumptions and limitations.

### Projected model outcomes

There could be unintended consequences of the options chosen, as these projections are for COVID-19 only and do not account for any interplay among other factors or diseases and their impact on vulnerable populations.

| *Scenario* | **New cases averted vs. baseline (%)** | **Deaths averted vs. baseline (%)** |
|---|---|---|
| ***Scenario 1*** | Reference | Reference |
| ***Scenario 2*** | +17.3 | +18.9 |
| ***Scenario 3*** | +28.6 | +48.4 |
| ***Scenario 4*** | +0.3 | +0.6 |
| ***Scenario 5*** | +33.9 | +53.6 |

The model predicted comparatively higher rates of new cases and deaths averted compared with the baseline in Scenarios 2 and 3 (lower and higher intensity managed lockdown releases). This includes the lockdown being lifted as planned on 10 May and followed up with:

- *self-isolation of symptomatic cases;*
- *case tracing;*
- *voluntary quarantine of those who had contact with COVID-19 positive cases*;
- *social distancing;*
- *hand hygiene and mask wearing.*

The 'highly socially disruptive scenario', with lockdown extended for 16 more weeks (scenario 5), was predicted to result in the highest percentage of averted new cases and deaths. However, this may result in adverse consequences for the social and economic life of the country and have unintended implications for people's mental health. It is interesting to note that the extension of the current lockdown without any follow-up interventions may have very little impact on the prevention of new cases and deaths (scenario 4).

Decisions on the strategy should be made with caution given the uncertainty around COVID-19 epidemiology (Graphs 1,2).

**Graph 1. Projected impact of intervention scenarios on COVID-19 cases.**

| | Incidence | Reproductive number ($R_t$)[1] | New cases averted |
|---|---|---|---|
| **Scenario 1 (Baseline)** | 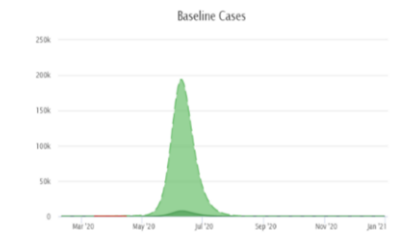 | 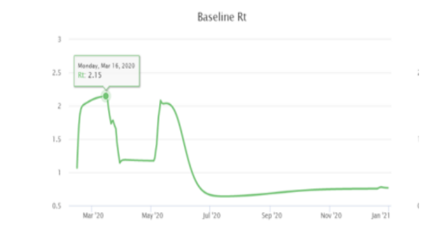 | Reference |
| **Scenario 2** | 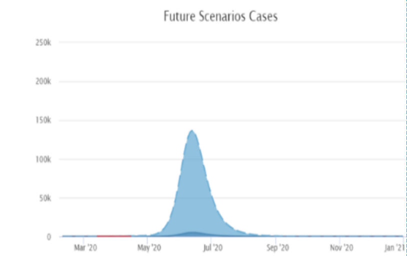 | 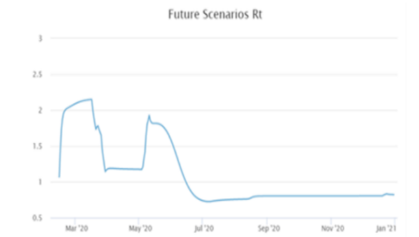 | 17.3 % |
| **Scenario 3** | 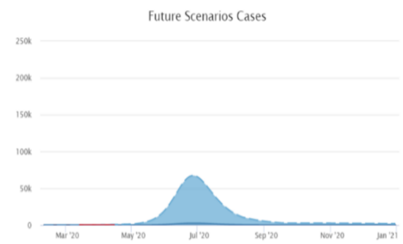 | 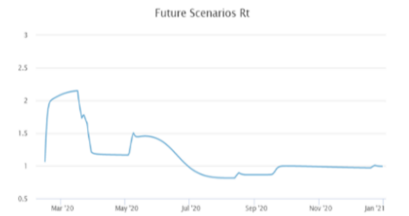 | 28.6% |
| **Scenario 4** | 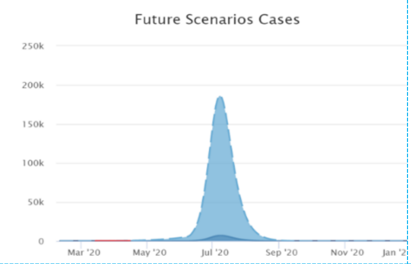 | 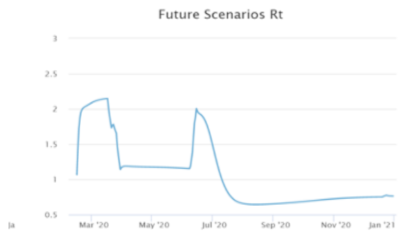 | 0.3% |
| **Scenario 5** | 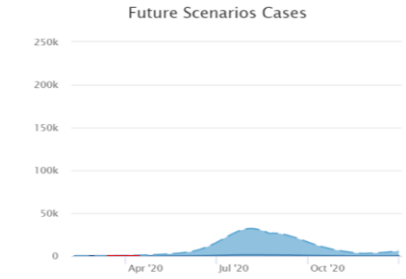 | 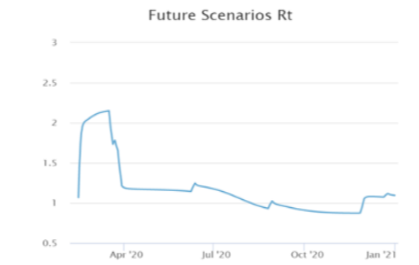 | 33.9% |

[1]***Note****: the reproductive number is an epidemiological value indicating the level of contagiousness of the infection, i.e. it is the expected number of cases generated by one infected person during the period of their disease. If Rt = 1, the epidemic is stabilised; if Rt > 1, the epidemic is increasing; if Rt < 1, the epidemic is decreasing.*

- In Scenario 1, the simulation predicts that the epidemic curve may sharply increase after the relaxation of the lockdown and that about 82.7% of the population may become infected in the ensuing months. It should be noted, however, that the majority of individuals may experience mild or no symptoms. Moreover, the reproductive number ($R_t$), currently balanced at the level of 1.2 to 1.3, may increase to about 2.1 immediately following the relaxation of lockdown then gradually decrease along with increasing immunity within the population as the virus is transmitted (i.e. achieving herd immunity). Note that Scenario 1 is referred as the 'baseline scenario' against which the other scenarios will be compared.

- In Scenario 2, the model predicts that, compared with the baseline scenario, the peak may decrease by around 20% and about 17.3% more new cases are likely to be averted. In total, about 65.0% of the population may get infected during the course of the epidemic (the majority with mild or no symptoms). Although an increase in the $R_t$ value after the relaxation of lockdown may still be observed, its trend may be slightly lower compared with the baseline scenario due to the extension of current interventions focussed on those who have symptoms or are diagnosed as COVID-19-positive and those who have had contact with positive cases.

- In Scenario 3, the model predicts that, compared with the baseline scenario, the peak may decrease by 70% and about 28.6% new cases are likely to be averted. In total, about 54.1% of the population may get infected during the course of the epidemic (the majority with mild or no symptoms). Although a slight increase in $R_t$ after the relaxation of lockdown may still be observed, it may decrease to 1.5, with a further decrease over time due to extended (19 weeks) and intensified interventions, focussed mainly on social distancing and those who have symptoms or are diagnosed positive and those who have had contact with positive cases.

- In Scenario 4, the model predicts that the peak may remain as high as in the baseline scenario, but the epidemic curve may move forward to a period roughly equivalent to the extension timeline (an additional 4 weeks). In total, about 82.4% of the population may get infected during the course of the epidemic (the majority with mild or no symptoms). The $R_t$ value may increase to the value equivalent to the baseline scenario and reach about 2.0 immediately after the relaxation of the lockdown, then gradually decrease as the virus is transmitted throughout the population (i.e. achieving herd immunity).

- Scenario 5 is likely to be the most effective in terms of epidemiological implications, but at the same time it includes highly disruptive interventions being extended for a longer time (an additional 8 weeks) after the initial lockdown. In this scenario, the model predicts that the peak will be flattened by 90% compared with the baseline option, and about 33.9% more new cases are likely to be averted. Moreover, the $R_t$ value will remain low, indicating a stabilised epidemic throughout the year. In total, about 48.8% of the population may get infected during the course of the epidemic (the majority with mild or no symptoms).

**Graph 2. Projected impact of intervention scenarios on cumulative deaths and projected requirements/needs for hospital and ICU beds and ventilation.**

<table>
<tr><th></th><th>Cumulative death</th><th>Hospital demand</th><th>Death cases averted</th></tr>
<tr><td>Scenario 1 (Baseline)</td><td>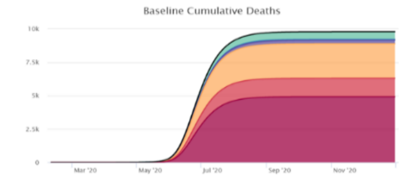
</td><td>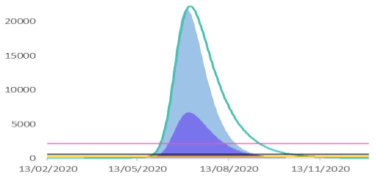
</td><td>Reference</td></tr>
<tr><td>Scenario 2</td><td>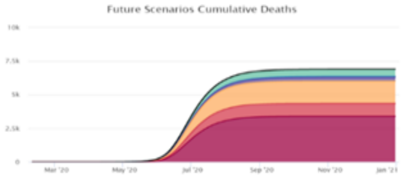
</td><td>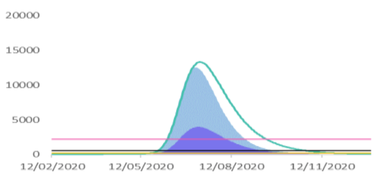
</td><td>18.9 %</td></tr>
<tr><td>Scenario 3</td><td>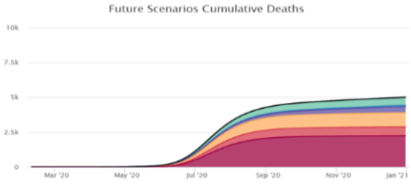
</td><td>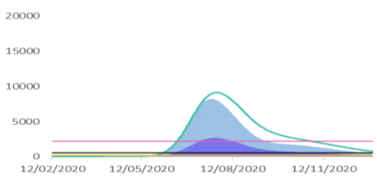
</td><td>48.4%</td></tr>
<tr><td>Scenario 4</td><td>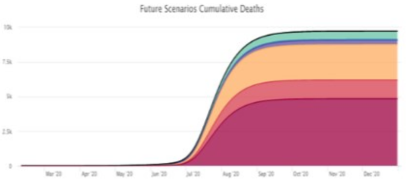
</td><td>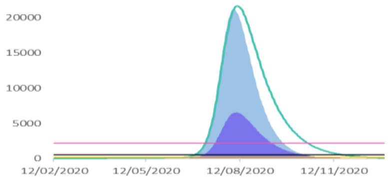
</td><td>0.6%</td></tr>
<tr><td>Scenario 5</td><td>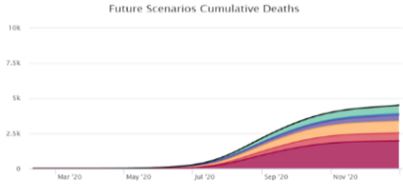
</td><td>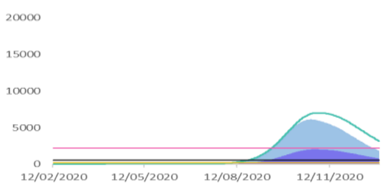
</td><td>53.6%</td></tr>
</table>

***Legend***

***Cumulative deaths among those who***

- required ventilation but were denied treatment
- required an ICU bed but were denied treatment
- required a hospital bed but were denied hospitalisation
- received ventilation treatment
- received ICU treatment
- received hospital treatment

***Hospital demand***

- required # of hospital beds
- required # of ICU beds
- required # of ventilators & oxygen equipment
- Surge beds capacity threshold
- ICU beds capacity threshold
- Ventilators capacity threshold

- In Scenario 1, the simulation predicts that, considering relevant interventions and planned hospital beds in general wards, ICU beds and ventilators, the health system will be overwhelmed with the influx of patients during the peak of the epidemic. As shown in Graph 2 ('hospital demand' column), the peak in the number of patients requiring medical assistance will significantly exceed the health system capacity (threshold lines), which will increase the possibility of denying necessary hospital support to many patients. As a result, the number of deaths among those who needed hospitalisation and intensive care support (including lung ventilation), but did not receive the necessary medical assistance, may become significant (refer to the 'cumulative death' column). Note that Scenario 1 is referred to as a 'baseline scenario', against which the other intervention options will be compared.

- In Scenario 2, the model predicts that the health system will still be highly overwhelmed by the influx of patients during the peak of the epidemic. However, the number of potential deaths among those who would be denied hospitalisation in the general ward or ICU or who would not be treated with lung ventilation will decrease slightly ('hospital requirements/needs' column). This may be due to the decrease in patients as a result of continued interventions focussed on limiting the spread of the infection from positive cases.

- In Scenario 3, the model predicts that the health system will be less burdened compared with the burden in the previous scenarios, although the number of surge and ICU beds and particularly ventilators will still be insufficient. This can be seen from the 'cumulative deaths' in Graph 2, where the number of people in need of ventilators but who have been denied necessary treatment is still high. The lower burden on the health system may be due to fewer patients as a result of continued and intensified interventions focussed on limiting the spread of the infection from positive cases.

- In Scenario 4, the simulation predicts that the health system will be overwhelmed at the same level as in the baseline scenario. As a result, the number of cases and deaths among those who need hospitalisation and intensive care support, including lung ventilation, but do not receive the necessary medical assistance, may become as significant, as in the baseline scenario.

- Scenario 5, as the simulation predicts, is the most effective in terms of epidemiological implications, with the least burden on the health system and the lowest number of deaths among those who might have been denied relevant hospital treatment. However, as mentioned earlier, this scenario involves the most disruptive interventions for a long period of time (an additional 8 weeks for lockdown plus other low disruptive interventions throughout the year), which is not feasible to implement in Kyrgyzstan.

##### Conclusion 1: Prevention is essential

It is critical to combine a phased transition after lockdown with key epidemiological prevention interventions, such as **quarantine** for those who have contact with positive cases, **self-isolation** if symptomatic, **screening** (testing), and **social distancing** to reduce transmission of the infection. **Hand hygiene and mask wearing** should remain an integral part of any set of measures. Rapid development of an effective, responsive and tailored risk communication strategy, including motivational strategies to encourage adherence to the key preventive interventions among the population, is important.

##### Conclusion 2: Rapidly scale-up screening and hospital capacities

**The model predicts the need for two approaches to be pursued simultaneously: prevention and health system preparedness.** A high level of prevention activities may reduce the burden on the health system, but the health system needs to ensure sufficient capacity to accept and treat all patients. Sufficient capacity in the health system for responding to increased demand in COVID-19 testing, contact tracing, quarantine and treatment are crucial for saving lives and reducing risks. A substantial focus on infection prevention control (IPC) is critical to address increasing rates of infected healthcare workers.

##### Conclusion 3: Uncertainty and updates

All results of the modelling should be used with caution, due to the limited evidence available about the spread of COVID-19 and its epidemiology. The model and its projections should constantly be updated as more evidence becomes available. It is also important to be aware of potential unintended consequences of the options chosen, as the model projections are for COVID-19 only and do not account for any interplay with other factors or diseases and the impact on general and vulnerable populations.

### Acknowledgements

We would like to thank the team and all of the contributors and organisations who helped with this project: Ministry of Health of the Kyrgyz Republic, Department of Disease Prevention and Surveillance under the Ministry of Health of the Kyrgyz Republic, Soros Foundation in the Kyrgyz Republic, Public Fund “Institution of Social Development” in the Kyrgyz Republic, and USAID Mission in the Kyrgyz Republic.

**Disclaimer**: The views expressed in this publication are those of the authors and do not necessarily reflect those of the Oxford Modelling Group for Global Health (OMGH), the COVID-19 Modelling Consortium (CoMo Consortium), the United States Agency for International Development, the United States Government, or the Soros Foundation of the Kyrgyz Republic and/or their funders.

### References


CDC China. (2020). *The Epidemiological Characteristics of an Outbreak of 2019 Novel Coronavirus Diseases (COVID-19) — China, 2020*. China CDC Weekly. https://doi.org/10.46234/CCDCW2020.032

EpiCOVID. (2020). *EpiCOVID: Round One*. https://ee.humanitarianresponse.info/x/#YKvCaTJf

Google Map. (2020). *COVID-19 Community Mobility Report*.

Korean Society of Infectious Diseases, Korean Society of Pediatric Infectious diseases, Korean Society of Epidemiology, Korean Society for Antimicrobial Therapy, Korean Society for Healthcare-associated Prevention, & Korea Centers for Disease Control and Infection Control and Prevention. (2020). Report on the Epidemiological Features of Coronavirus Disease 2019 (COVID-19) Outbreak in the Republic of Korea from January 19 to March 2, 2020. *Journal of Korean Medical Science*, *35*(10). https://doi.org/10.3346/jkms.2020.35.e112

Liu, T., Hu, J., Kang, M., Lin, L., Zhong, H., Xiao, J., He, G., Song, T., Huang, Q., Rong, Z., Deng, A., Zeng, W., Tan, X., Zeng, S., Zhu, Z., Li, J., Wan, D., Lu, J., Deng, H., … Ma, W. (2020). Transmission dynamics of 2019 novel coronavirus (2019-nCoV). *BioRxiv*, 2020.01.25.919787. https://doi.org/10.1101/2020.01.25.919787

Prem, K., Cook, A. R., & Jit, M. (2017). Projecting social contact matrices in 152 countries using contact surveys and demographic data. *PLoS Computational Biology*, *13*(9), e1005697. https://doi.org/10.1371/journal.pcbi.1005697

Riou, J., Hauser, A., Counotte, M. J., & Althaus, C. L. (2020). Adjusted Age-Specific Case Fatality Ratio During the COVID-19 Epidemic in Hubei, China. *MedRxiv PrePrint*. https://doi.org/10.1101/2020.03.04.20031104

UN. (2020). *World Population Prospects - Population Division - United Nations*. https://population.un.org/wpp/Download/Standard/Population/

WHO. (2020a). *COVID19 Situation reports*. https://www.who.int/emergencies/diseases/novel-coronavirus-2019/situation-reports/

WHO. (2020b). *Health Systems Respond to COVID-19 Technical Guidance #2 Creating surge capacity for acute and intensive care Recommendations for the WHO European Region (6 April 2020)*. http://www.euro.who.int/__data/assets/pdf_file/0006/437469/TG2-CreatingSurgeAcuteICUcapacity-eng.pdf

MoH. (2020a). *Daily COVID-19 situation report as of April 24, 2020*

MoH. *(2020b). MoH report on health system preparedness for the COVID-19 epidemic.*

DRCU.*(2020) Weekly update of Disaster Response Coordination Unit in the Kyrgyz Republic, as of 17 April, 2020*
